# The implementation of liquid biopsies in routine care of patients with advanced solid cancer (LIQPLAT): a study protocol for a single arm trial

**DOI:** 10.1101/2025.02.13.25322206

**Authors:** Johannes M. Schwenke, Andreas M. Schmitt, Perrine Janiaud, Heinz Läubli, Mascha Binder, Ilaria Alborelli, Matthias S. Matter, Jennifer Hinke, Corinne C. Widmer, Lars G. Hemkens, Benjamin Kasenda

## Abstract

- **Background:** Advanced solid cancers present significant treatment challenges due to their genomic heterogeneity and resistance. Liquid biopsies, specifically circulating tumour DNA (ctDNA), have emerged as promising tools to support treatment decision making. However, evidence regarding their implementation in routine care remains limited.
- **Methods:** LIQPLAT is a single-arm trial (SAT), assessing the feasibility of implementing ctDNA measurements in the usual care of patients with advanced solid cancers excepting primary brain tumours, at the University Hospital Basel. Patients are randomly invited from an ongoing research registry to take part in the SAT. We aim to include 150 randomly invited patients to receive ctDNA measurements alongside standard care. CtDNA samples are collected at baseline, between the second and third months, fifth and seventh months after cancer treatment start, and during serious clinical events such as disease progression or treatment changes. Results are evaluated by a molecular tumor board to guide clinical management. Feasibility outcomes include detectability of ctDNA, identification of actionable alterations, and analysis turnaround time. Other outcomes include patient-reported quality of life, progression-free survival and overall survival, time to next treatment line, and unscheduled hospital and emergency visits, all obtained from routine healthcare data.
- **Discussion:** LIQPLAT will examine the implementation of ctDNA measurements in routine care. The random selection for invitation to this SAT within an existing registry embedded in routine care creates a representative sample and allows for better assessment of implementation and generalization of findings.
- **Trial registration:** The trial is registered at clinicaltrials.gov (2024-04-11, NCT06367751) and kofam.ch (2024-03-15, SNCTP000005844).

## Administrative information

Note: the numbers in curly brackets in this protocol refer to SPIRIT checklist item numbers. The order of the items has been modified to group similar items (see http://www.equator-network.org/reporting-guidelines/spirit-2013-statement-defining-standard-protocol-items-for-clinical-trials/).

|  |  |
| --- | --- |
| Title {1} | The implementation of liquid biopsies in routine care of patients with advanced solid cancer (LIQPLAT): a study protocol for a single arm trial |
| Trial registration {2a and 2b}. | The trial is registered at clinicaltrials.gov (2024-04-11, NCT06367751) and kofam.ch (2024-03-15, SNCTP000005844). |
| Protocol version {3} | Version 1.1 (dated 2024-03-14) |
| Funding {4} | This trial is currently being funded by the *Krebsliga beider Basel* (KLbB-5845-02-2023) and the Department of Medical Oncology. The *Krebsliga beider Basel* has no role in any aspect of the study or writing of the manuscript. JMS is funded by the Swiss National Science Foundation (MD-PhD grant number: 323530_229969). |
| Author details {5a} | <sup>1</sup> CLEAR Methods Center, Division of Clinical Epidemiology, Department of Clinical Research, University Hospital Basel, University of Basel, Basel, Switzerland<br><sup>2</sup> Division of Medical Oncology, University and University Hospital Basel, Basel, Switzerland<br><sup>3</sup> Pragmatic Evidence Lab, Research Center for Clinical Neuroimmunology and Neuroscience Basel (RC2NB), University Hospital Basel and University of Basel, Basel, Switzerland.<br><sup>4</sup> Meta-Research Innovation Center at Stanford (METRICS), Stanford University, Stanford, CA, USA<br><sup>5</sup> Laboratory of Translational Immuno-Oncology, Department of |
|  | Biomedicine, University and University Hospital Basel, Basel, Switzerland.<br><sup>6</sup> Department of Pathology, Institute of Medical Genetics and Pathology, University Hospital Basel, University of Basel, Basel, Switzerland.<br><sup>7</sup> Division of Hematology, University Hospital Basel, Basel, Switzerland.<br>* These authors contributed equally.<br># These authors are shared last authors.<br>† Corresponding author: |
| Name and contact information for the trial sponsor {5b} | Sponsor: University Hospital Basel, Petersgraben 4, 4031 Basel<br>Sponsor investigator: Benjamin Kasenda |
| Role of sponsor {5c} | The University Hospital Basel serves as the sponsor for this study. The sponsor provides institutional oversight and administrative resources. However, the sponsor does not directly influence the study design, data collection, analysis, interpretation, or the reporting and publication of results. |

## Introduction

### Background and rationale {6a}

One of the major challenges in treating patients with solid cancers is the cancer’s high genomic heterogeneity and the consecutive resistance to cancer treatments (1). Liquid biopsies have been proposed as a potential tool to help address these challenges. Liquid biopsies encompass the detection and analysis of nucleic acids, tumour cells, or exosomes in body fluids, including peripheral blood, urine, or cerebrospinal fluid (2–4). While circulating biomarkers are well-established in haematology, there has been little uptake in medical oncology. Nevertheless, technologies to analyse circulating tumour DNA (ctDNA) in liquid biopsies have rapidly evolved over recent years, and more and more researchers advocate for their use in clinical care (5).

Measuring ctDNA may help to (i) *identify targetable alterations* due to improved genomic representation of tumour heterogeneity compared to solid biopsy only (6–8). Especially in solid cancers, the molecular pattern derived from a single biopsy is often limited to a single anatomical site and to a specific point in time, not reflecting tumour heterogeneity. In contrast, ctDNA may better map the cancer’s genomic anatomical and chronological variability. Furthermore, ctDNA can help to (ii) *quantify disease burden and response to treatment* (9,10), (iii) *identify early mutations of resistance*, which would allow for prompt adaptations of therapy (11), and (iv) *improve prediction of outcomes* (12,13). This contrasts with conventional biomarkers, such as CA19-9 or prostate specific antigen, that can provide information on disease activity and treatment response but may only provide limited information on cancer biology and cannot identify targetable alterations or resistance mechanisms.

Analysing ctDNA presents challenges, including ‘background noise’ from Clonal Haematopoiesis of Indeterminate Potential (CHIP), an age-related somatic mutation process observed mostly in patients over 70, which can complicate data interpretation (14–16). Although bioinformatic approaches can be applied to filter known CHIP alterations, the impact of CHIP on the use of ctDNA in routine care for advanced cancer patients in unknown.

The increasing availability of standardized technologies to detect ctDNA has led to first recommendations for the use of ctDNA measurements in clinical practice (5), but with very little evidence from randomized trials, especially in patients with advanced / metastatic malignancies. Therefore, there is a need to further investigate its implementation in routine care of patients with advanced cancers (17). Here we describe the design of a single-arm trial (SAT) to investigate ctDNA implementation into routine care in a representative sample of patients with advanced cancers. To facilitate representative sampling of the local target population and enhance the applicability of findings, the trial participants are randomly invited to join the trial.

### Objectives {7}

The primary objective of this trial is to assess the implementation and feasibility of repeated ctDNA measurements from peripheral blood during routine clinical care of cancer patients in the University Hospital Basel, Switzerland.

### Trial design {8}

LIQPLAT is a single-centre SAT embedded in an ongoing prospective research registry at the University Hospital of Basel. Healthcare data of patients are routinely collected in the electronic health record system and mirrored in the hospital’s clinical data warehouse (CDWH) and REDCap databases (18,19) of the Division of Medical Oncology. Patients who have signed the hospital’s General Research Consent (GRC) (20,21) form a prospective registry and their data are available for further use for research. Eligible participants from the registry are randomly invited to take part in LIQPLAT (***Figure 1***). The trial almost exclusively uses routinely collected data. LIQPLAT aims to generate evidence in an environment as close as possible to routine care (22).

**Figure 1.**
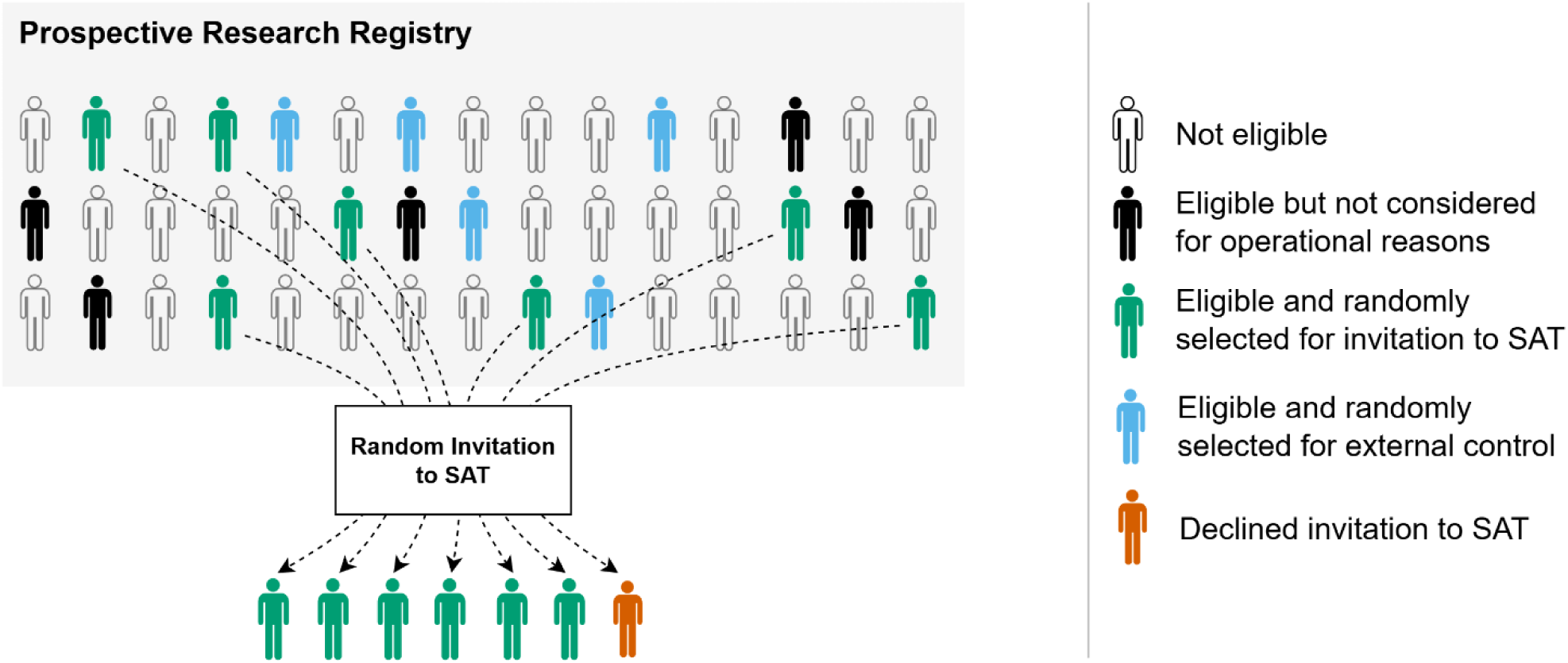
Trial design. All patients of the University Hospital Basel who have signed the general research consent form a prospective research registry. From this population, eligible patients are randomly invited to the SAT. Some eligible participants might not be considered for invitation for operational reasons, e.g., inpatients who require emergency treatment before trial staff is made aware of their existence. Patients who were considered for invitation, but not randomly selected, are flagged as potential, registry-based controls. ***Abbreviations***: SAT, single-arm trial.

## Methods: Participants, interventions and outcomes

### Study setting {9}

The trial is fully embedded in the routine care of the University Hospital Basel’s Division of Medical Oncology. Usual care for advanced cancer patients consists of regular visits, which include clinical assessment, blood samples, assessment of treatment response, and regular imaging studies every 2 to 4 months. The exact schedule is at the physician’s and patient’s discretion and may thus vary over time depending on treatment and disease situation.

### Eligibility criteria {10}

We are aiming to generate evidence for all patients with advanced solid cancers who might receive ctDNA measurements in real-world care settings. We therefore avoid eligibility criteria that would unnecessarily exclude members of the target population.

We include:

1. all adult patients who are part of the University Hospital Basel’s prospective research registry, i.e., have signed the GRC, with
2. a proven advanced solid malignant disease and
3. an indication for medical anti-cancer treatment (including combined chemo-radiotherapy or immuno-radiotherapy).

We exclude:

1. patients with primary brain tumours, as these patients do not present with or only very low amount of ctDNA in plasma (23),
2. patients with primary resectable disease, and
3. patients with prior treatment for advanced or metastatic disease, as ctDNA dynamics between a measurement prior to treatment and an early on-treatment timepoint are likely crucial for assessing molecular response (10).

### Who will take informed consent? {26a}

The treating oncologist or a trial physician offers eligible patients, that were randomly selected, participation in the SAT. This invitation is prior to treatment start, during an outpatient usual care visit to the Division of Medical Oncology or during a hospitalization for their underlying condition. All participants provide written informed consent in addition to the GRC they have already provided.

### Additional consent provisions for collection and use of participant data and biological specimens {26b}

n/a. All participants have provided the GRC.

## Interventions

### Explanation for the choice of comparators {6b}

n/a.

### Intervention description {11a}

All patients who agree to participate in the SAT receive routine care disease assessment and treatment, plus a blood draw for ctDNA measurement at specific time points or events:

1. Before start of medical anticancer treatment.
2. Between the second and third months after treatment start; typically, after a first treatment block.
3. Between the fifth and seventh month of treatment.
4. At any event of suspected or confirmed clinical or radiological disease progression or treatment discontinuation for any reason.

We use existing routine workflows that are established at the Department of Pathology to process and analyse ctDNA. We use the Oncomine™ Pan-Cancer Cell-Free Assay (24) from Thermo Fisher Scientific to analyse ctDNA for all participants except for participants with prostate cancer, where a custom prostate cancer panel (25) is used.

The Pan-Cancer Cell-Free Assay covers the following alterations. DNA hotspots: *AKT1, ALK, AR, ARAF, BRAF, CHEK2, CTNNB1, DDR2, EGFR, ERBB2, ERBB3, ESR1, FGFR1, FGFR2, FGFR3, FGFR4, FLT3, GNA11, GNAQ, GNAS, HRAS, IDH1, IDH2, KIT, KRAS, MAP2K1, MAP2K2, MET, MTOR, NRAS, NTRK1, NTRK3, PDGFRA, PIK3CA, RAF1, RET, ROS1, SF3B1, SMAD4, SMO.* Tumour suppressors: *APC, FBXW7, PTEN, TP53.* Copy number variations: *CCND1, CCND2, CCND3, CDK4, CDK6, EGFR, ERBB2, FGFR1, FGFR2, FGFR3, MET, MYC. Fusions: ALK, BRAF, ERG, ETV1, FGFR1, FGFR2, FGFR3, MET, NTRK1, NTRK3, RET, ROS1*.

The custom prostate cancer panel covers the following alterations: *ACVR2A, AKT1, APC, AR, ASXL1, ATM, BRAF, BRCA1, BRCA2, CDK12, CHD1, CHEK2, CSMD3, CTNNB1, CUL3, CYP17A1, CYP19A1, FBXW7, FOXA1, HRAS, HSD17B4, HSD3B1, IDH1, JAK1, KDM6A, KMT2C, KRAS, LHCGR, MED12, MGA, MYC, NCOR1, NCOR2, PIK3CA, PIK3R1, PTEN, RB1, RNF43, SLCO1B1, SLCO1B3, SPOP, TP53, ZBTB16, ZFHX3, ZMYM3, ZNF780B*.

The input cell-free DNA used for library preparation is 20 ng, if available, which theoretically allows to obtain a limit of detection down to 0.1% molecular frequency.

CtDNA measurements may influence the clinical management of participants through the molecular tumour board (MTB). The MTB is an established interdisciplinary meeting at the University Hospital Basel to discuss patients with complex findings from tumour sequencing analyses. Results from ctDNA analyses are discussed at the MTB and each discussion is summarised in a recommendation for the responsible clinical team. We always try to match ctDNA results with next generation sequency (NGS) results based on solid tumour tissue when available from routine care. We will primarily use the allelic frequency to quantify and compare the ctDNA kinetics over time. Because of the heterogeneity of diseases and associated treatment histories, we have not pre-defined exact recommendations on how to further individualize clinical management based on ctDNA results.

### Criteria for discontinuing or modifying allocated interventions {11b}

Participants may withdraw their consent and discontinue participation in the trial at any time. Following withdrawal, they will continue to receive usual care. If a participant withdraws the GRC during the study, we will not use their routinely collected data for the analysis from that timepoint on.

### Strategies to improve adherence to interventions {11c}

No specific measures are implemented to increase adherence to ctDNA measurements.

### Relevant concomitant care permitted or prohibited during the trial {11d}

No participant included in the trial will be denied anything for the purpose of research or will have to do anything extra solely for the purpose of research. All participants can also be included in other research projects, including clinical trials if they wish.

### Provisions for post-trial care {30}

There is no specific post-trial care beyond usual care.

### Outcomes {12}

#### Feasibility outcomes

The feasibility outcomes are: the number and proportion of participants in whom ctDNA was detectable before starting medical anticancer treatment, the number and proportion of participants in whom actionable alterations were identified in ctDNA analysis, the number and proportion of ctDNA analyses carried out without error, the number and proportion of ctDNA analyses being labelled with suspicion of CHIP, the turnaround time of ctDNA, ctDNA kinetics over time including relative changes from baseline expressed as changes in variant allele frequency, and the number and proportion of patients who accept the invitation to the trial.

#### Further outcomes

We will also describe clinical endpoints, including overall and progression-free survival, time to next treatment line, and the frequency of unplanned hospital admissions and emergency room visits of trial participants, as well as evolution of patient reported global quality of life (QoL) and physical functioning according to European Organisation for Research and Treatment of Cancer (EORTC) questionnaires over time. We will also assess participant referral to other clinical trials, the frequency and proportion of tissue biopsies, the number of imaging studies performed per participant, the number of blood product transfusions (red blood cells or platelets), and the cumulative doses of cancer treatments administered.

### Participant timeline {13}

We anticipate a recruitment period of 18 months, which may be adapted depending on recruitment rates. As all participants will have their follow-up as per routine care, the length of follow-up is not limited by the trial. Minimum follow-up is 12 months (see ***Table 1***), so that we anticipate a trial duration of 30 months in total.

**Table 1.** Study assessment. *While ctDNA measurements are not established as part of routine care, all data can be extracted from established, routine processes. **Abbreviations**: ctDNA, circulating tumour DNA; EORTC-QLQ, EORTC Core Quality of Life questionnaire; CT, computed tomography; PET, Positron emission tomography; MRI, magnetic resonance imaging.

|  | Pre-Baseline | Baseline | Follow-up |  |  |
| --- | --- | --- | --- | --- | --- |
| Time |  | M0 | M2-3 | M5-7 | M12 |
| Recruitment |  |  |  |  |  |
| Eligibility screening | X |  |  |  |  |
| Informed consent |  | X |  |  |  |
| Random selection | X |  |  |  |  |
| Baseline characteristics | X | X |  |  |  |
| Intervention |  |  |  |  |  |
| Routine ctDNA sampling |  | X | X | X |  |
| ctDNA sampling if (suspected) disease progression or toxicity |  |  | At any point |  |  |
| Endpoints (all gathered from routinely collected data and routine processes) |  |  |  |  |  |
| ctDNA feasibility |  |  |  |  |  |
| Detectable ctDNA at baseline* |  | X |  |  |  |
| Actionable alterations identified on ctDNA* |  | X | X | X |  |
| Turn-around time of ctDNA* |  | X | X | X |  |
| ctDNA kinetics* |  | X | X | X |  |
| Quality of life |  |  |  |  |  |
| EORTC-QLQ C30 |  | X |  |  |  |
| EORTC-QLQ C15 |  |  | X | X | X |
| Clinical endpoints |  |  |  |  |  |
| Overall survival |  |  |  | X | X |
| Progression free survival |  |  |  | X | X |
| Time to next treatment line |  |  |  | X | X |
| Unplanned hospital admissions and emergency room visits |  |  |  | X | X |
| Further endpoints |  |  |  |  |  |
| Referral to different interventional clinical trial |  |  |  | X | X |
| Number of solid tissue biopsies and proportion of patients receiving a biopsy |  |  |  | X | X |
| Number of imaging exams per patient (CT, PET/CT or MRI) |  |  |  | X | X |
| Number of blood products per patient |  |  |  | X | X |
| Cumulative doses of cancer drug treatments administered |  |  |  | X | X |

### Sample size {14}

Based on past-data of the Division of Medical Oncology, around 240-300 patients would be eligible for the trial over a period of 18 months. However, for logistic reasons and capacity of the Department of Pathology, we limit the sample size to 150 participants, with a maximum of three patients selected for invitation to the trial per week.

### Recruitment {15}

We aim to include 150 participants who accept the invitation to the trial over 18 months. For example, if 5% of patients invited to the trial did not wish to participate, we would need to invite at least 158 patients. We anticipate a very high up-take of the invitation due to the expected good integration into routine practice, as ctDNA sampling requires no extra effort from the patients and is performed at the same time as a routine venipuncture, avoiding additional discomfort.

### Assignment of interventions: allocation {16}

n/a, as it is a single arm trial.

### Assignment of interventions: Blinding {17}

n/a, as it is a single arm trial.

### Random invitation

#### Screening list

We build a weekly screening list containing all patients who have an appointment for a first consultation at the Division of Medical Oncology the following week in REDCap (v. 14.3.14), using routinely collected data from the electronic patient records. The list is randomly ordered by sorting by a hashed version (SHA-256) of the sequentially generated patient hospital ID. The trial team (at least one oncologist) checks whether patients fulfil the eligibility criteria every Friday afternoon.

Invitation to SAT.

Descending the screening list row by row, we select up to three patients per week who will be invited to the trial. The decision to invite is made randomly, using a random sampling table uploaded to REDCap. The random sampling table was generated by a trialist not associated with the trial using R version 4.3.1 (2023-06-16). The table was stratified by presence or absence of a primary diagnosis of a lung tumour, with a block size of 9, and has odds of 2:1 for an invitation to the trial. All investigators are blinded to the random sampling table. Once three patients have been randomly selected for an invitation to the trial, the list is closed for the current week and a new one is started the next week. Inpatients can also be invited when fulfilling eligibility criteria and slots are available for the given week.

## Data collection and management

### Plans for assessment and collection of outcomes {18a}

We exclusively use routinely collected data for outcome measures (see ***Table 1***). Since October 2022, the Division of Medical Oncology has been documenting and coding all cancer diagnoses using the *OncoTree* nomenclature (26) and clinical metrics such as ECOG performance status in a validated REDCap database integrated into the hospital information system.

Since June 2023, as part of usual care, patients with a first encounter are invited to complete the EORTC QLQ-C30, with response rates of about 90% so far. For regular follow-up, the shorter EORTC QLQ-C15 is used to maintain patient engagement. Similar to the baseline questionnaire, about 90% of questionnaires handed out are completed. All questionnaires are answered on tablets and managed with REDCap.

Clinical outcomes such as survival, hospitalization, imaging and laboratory results are recorded in the hospital’s CDWH, which also receives regular updates from administrative databases.

### Plans to promote participant retention and complete follow-up {18b}

No specific measures are implemented to increase patient retention for the purpose of the trial as this would go against the aim of assessing the implementation in a routine care setting.

### Data management {19}

The REDCap database is hosted on the servers of the University Hospital of Basel. All data entries and modifications are recorded in an audit trail. Password protection and user-right management ensure that only authorized personnel have access to the data. For all analyses, we will use the software R to pull data from the REDCap database and the CDWH. The unique patient hospital ID will be the linker throughout all merging of data tables. All data will remain on the servers of the University Hospital Basel and only dedicated trial personnel will have access to the merged dataset. A dedicated and version-controlled data management plan is being implemented in a GitLab repository hosted on the servers of the University Hospital Basel. In case of data sharing with investigators outside the University Hospital Basel, we will follow the guidelines of Swiss Personalized Health Network (SPHN) for data sharing and de-identification (27).

### Confidentiality {27}

Trial and participant data will be handled as routine clinical care data; thus, all data is only accessible to authorized personnel who are involved in the care of the patients and study personnel to fulfil their duties within the scope of the study. Specifically, all persons involved in the conduct of the trial must be employees of the University Hospital Basel and have signed the standard data protection agreements applicable to all staff being involved in patient care. When consulting with trial personnel not involved in patient care, e.g., the trial statistician, all data will be pseudonymized.

### Plans for collection, laboratory evaluation and storage of biological specimens for genetic or molecular analysis in this trial/future use {33}

The site will retain all essential documents according to ICH GCP and all applicable Swiss laws and regulations. This includes participant trial records, which are considered as source data, electronic case report forms, participant informed consent statement, and all other information collected during the trial. These documents will be stored for at least 20 years after the last participant’s last visit (LPLV) in the trial. For the participant trial records, which are entered into the electronic data capture system, the sponsor guarantees the access and availability of the data at any time at least 20 years after the LPLV of the trial. In the event that the principal investigator retires or changes employment, custody of the records may be transferred to another competent person who will accept responsibility for those records. The sponsor will notify the concerned regulatory authorities.

## Statistical methods

### Statistical methods for primary and secondary outcomes {20a}

Because the primary aim of this trial is assessment of feasibility, we will conduct descriptive analysis on the pre-specified endpoints and potentially additional endpoints if deemed necessary: median/mean, and range for continuous data; frequencies/proportions with 95% confidence intervals for categorical data; frequency and proportion of missing data items.

### Interim analyses {21b}

n/a. No interim analyses are planned for this study.

### Methods for additional analyses (e.g. subgroup analyses) {20b}

We will conduct the above-mentioned descriptive analyses also stratified by the different cancer indications.

### Methods in analysis to handle protocol non-adherence and any statistical methods to handle missing data {20c}

We will report the frequency of missing data for all relevant variables. For routinely collected patient reported outcomes measures (PROMs) at the Division of Medical Oncology we have so far observed response rates of about 90%. Thus, the occurrence of missing data is expected to be modest for QoL metrics. Clinical outcome-related data also stems from routinely collected data, for which we expect low missingness.

### Plans to give access to the full protocol, participant level-data and statistical code {31c}

Statistical code will be made available on GitHub. Anonymized participant-level data will be made available upon reasonable request to the principal investigator.

### Process Evaluation

To better understand the implementation of the ctDNA in routine practice, which constitutes a complex intervention, an embedded process evaluation will further explore the implementation and impact of ctDNA measurement in routine clinical patient care. Process evaluations enhance our understanding of why interventions succeed or fail, by examining factors such as intervention delivery, uptake, and contextual factors (28,29). We explain the process evaluation in detail separately in an upcoming publication but outline the approach in brief below.

We developed a logic model for the process evaluation and mapped this logic model to the RE-AIM framework (Reach, Effectiveness, Adoption, Implementation, Maintenance) (30,31). The process evaluation uses a mixed-methods approach to examine delivery, adoption, implementation, as well as contextual factors of the complex intervention. Key aspects include assessing how ctDNA measurements influence clinical decisions, identifying barriers and facilitators encountered by healthcare providers, and quantifying the extent to which the intervention was fully implemented. The evaluation uses detailed qualitative analyses, such as thematic analysis (32) of interviews with stakeholders—including participants, pathologists, and oncologists—and analysis of molecular tumour board session recordings. Quantitatively, it will examine assumptions of the logic model, such as number of targeted therapies delivered or time to treatment modification.

## Oversight and monitoring

### Composition of the coordinating centre and trial steering committee {5d}

The day-to-day operations of the trial are managed by a core team comprising: a consultant oncologist, a senior consultant oncologist, a medical doctor pursuing a PhD in clinical epidemiology, and a trial coordinator. This team meets weekly on Fridays to screen the next week’s patients. In addition, there are regular meetings with three trial methodologists and staff from the of department of molecular pathology to discuss trial progress, issues, and make key decisions. BK is the legal sponsor investigator (principal investigator) holding responsibility of the trial.

### Composition of the data monitoring committee, its role and reporting structure {21a}

n/a. LIQPLAT is a low-risk trial, fully embedded in usual care.

### Adverse event reporting and harms {22}

We do not consider specific safety reporting in this trial, because there is no trial procedure that puts the participant at harm, e.g., no trial specific drug intervention or invasive diagnostic intervention. The only interventions required are peripheral venous blood draws to measure ctDNA. Patients receiving treatment for advanced malignancies receive regular (sometimes weekly) venous blood samples, either by peripheral venous puncture or by blood sample from existing devices with central venous catheters (e.g., port-a-cath systems). We will take the additional sample for ctDNA analysis at the same time as routine blood testing under treatment. Therefore, there is no additional safety reporting in terms of serious adverse reactions (SARs) as necessary beyond standard clinical practice.

### Frequency and plans for auditing trial conduct {23}

n/a. LIQPLAT is a low-risk trial, fully embedded in routine patient care.

### Plans for communicating important protocol amendments to relevant parties (e.g. trial participants, ethical committees) {25}

Amendments will be signed by all signatories of the protocol. All investigators will acknowledge the receipt and confirm by their signature that they will adhere to the amendment. The principal investigator will be responsible for implementing any amendments at the study site (including the distribution of amendments to all staff concerned).

### Dissemination plans {31a}

Results from this trial will be made available via poster and/or oral presentations at national and international oncology congresses and via publication in peer-reviewed medical journals - whenever possible with open access. Results will also be made available on preprint servers and via the SPHN network infrastructure in Switzerland.

## Discussion

Cancer remains the leading cause of death and lost life years in Switzerland (33), despite improved treatment and public health efforts (34). Here we present the design and rationale of a single-arm trial to assess the implementation of routine ctDNA measurements in patients with advanced solid cancer, embedded within a prospective research registry. Given resource constraints, particularly of the Department of Pathology carrying out the ctDNA analyses, we would not have been able to offer routine ctDNA measurements to all advanced cancer patients. We thus opted for an approach which ensures feasibility and fairness, through random invitation of up to three patients per week from the ongoing research registry to take part in the trial.

The study design is highly practical and leverages the availability of routinely collected data to optimally embed the trial in routine care. This allows for a resource-saving approach and a low burden for the clinical teams and patients, which is crucial to achieve high acceptability and implementation into routine clinical workflows.

Random selection of patients for study invitation aligns with our patient-centered approach as current evidence does not clearly indicate which groups of advanced cancer patients would benefit most from routine ctDNA measurements. Patients not selected for the study will receive the same care they would have received if the SAT was not taking place, including the possibility of ctDNA analysis if deemed necessary by their physician, though we expect this to be very rare. All participants of the SAT can also be included in any other research project, including clinical trials, if they wish.

The Division of Medical Oncology at the University Hospital Basel serves a large metropolitan catchment area in Northwestern Switzerland, treating cancer patients with various malignancies. It is not only a tertiary referral centre offering all sorts of clinical trials (including phase I trials), but also the largest caregiver for cancer patients in Northwestern Switzerland. Thus, we can assume that the patient population being treated at the Medical Oncology Division is representative of the great majority of cancer patients. Keeping this in mind, the systematic random sampling approach, together with the broad inclusion criteria, should guarantee a representative sample of advanced cancer patients being recruited for the LIQPLAT trial, provided they have consented to the GRC, which is the case in more than 80%. However, we cannot exclude the possibility that patients who consent to the GRC are systematically different from patients who do not wish to have their routine data used for clinical research. This issue has not been investigated so far.

Random invitation allows for the estimation of valid uncertainty estimates for inferences derived from the sample and extrapolation to the larger population of cancer patients fulfilling the inclusion criteria. This estimation is difficult in most clinical trials, as these nearly always use convenience sampling with strict eligibility criteria (35,36). It has been highlighted by regulators that external validity is often compromised in single arm trials (37). Our approach enhances the generalizability and applicability of our results, which is crucial for implementation studies that aim to be directly translated into usual care.

Our random invitation process provides very high yet not perfect representativeness of the target population. This is because outpatients are considered during weekly Friday screenings, with a maximum of three selected. Inpatients, such as emergency admissions for newly diagnosed cancers requiring immediate treatment obviously cannot be included in these screenings. They are only invited if fewer than three outpatients were selected the previous Friday, leading to inpatients being less frequently included overall.

Similarly, outpatients meeting all criteria except the GRC remain on the screening list, unless three patients have already been selected that week. If a patient signs the GRC during their first contact with the Division of Medical Oncology, they may then be invited to the trial. This approach favours outpatients and those who have already signed the GRC. Consequently, our sample may be less representative of acutely ill inpatients (e.g., patients with newly diagnosed and symptomatic small cell lung cancer), which may affect some validity of sample-to-population inferences. However, the great majority of patients with advanced cancers (over 80%) start treatment in an outpatient setting including the frail and those with poor performance status.

Of note, as the decision to invite a patient to the trial is random, and we further know which patients of the research registry were not chosen for invitation, we are able to compare outcomes between trial participants and other patients of the research registry, i.e., an external control group, without confounding at baseline. This approach will be described in more detail elsewhere.

We use the commercially available Oncomine™ Pan-Cancer Cell-Free Assay (Thermo Fisher) for the LIQPLAT trial. This assay covers 52 genes including relevant hotspot genes (single nucleotide variants) and short indels, gene fusions, and copy number variations (24). We have decided to use this panel, because it is well established and validated at the Division of Molecular Pathology at the University Hospital Basel and covers most alterations that have actionable potential. We are aware of larger panels, however, the additional benefit of larger, and more costly panels to guide decision making is unclear. Thus, we believe that the 52-gene panel is a sound basis for the objectives of the LIQPLAT trial. For patients with prostate cancer we opted for a custom panel (Thermo Fisher). This panel is also established at the Division of Molecular Pathology and covers treatment relevant alterations for prostate cancer not covered by the 52-gene panel, such as *BRCA1* and *BRCA2.* All results from ctDNA are discussed at the regular interdisciplinary MTB. Because of the heterogeneity of malignancies included, we have decided against exact guidelines on how to translate ctDNA findings into treatment recommendations. We will always assess and discuss the findings in the context of the entire disease and treatment history. We anticipate that recommendations from the MTB will mostly be implemented as suggested, especially in situations with clinical uncertainty.

In conclusion, the LIQPLAT trial represents a significant step forward in evaluating the feasibility of integrating routine ctDNA measurements into clinical practice for patients with advanced solid cancers. By embedding our project within routine care processes and structures, this study aims to provide first, generalizable data on the implementation of ctDNA analyses. The trial’s innovative design ensures fair participant selection, optimizes resource use, and allows for secondary comparative effectiveness analysis with non-invited patients of the research registry. Ultimately, LIQPLAT seeks to lay the groundwork for larger trials, possibly paving the way for broader implementation of personalized cancer care based on ctDNA insights.

### Trial status

The first patient was randomly selected for invitation to the trial on April 29^th^ 2024, the first patient was invited on May 2^nd^ 2024. As of September 20^th^ 2024 48 patients have been selection for invitation to the SAT, of which 41 have accepted the invitation. Recruitment is scheduled to be completed in October 2025.

## Abbreviations

- **BASEC**: Business Administration System for Ethics Committees
- **CDWH**: Clinical Data Warehouse
- **CHIP**: Clonal Haematopoiesis of Indeterminate Potential
- **ctDNA**: Circulating Tumour DNA
- **EORTC**: European Organisation for Research and Treatment of Cancer
- **GCP**: Good Clinical Practice
- **GRC**: General Research Consent
- **ICH**: International Conference on Harmonisation
- **LIQPLAT**: Liquid biopsies in routine care of patients with advanced solid cancer
- **LPLV**: Last Patient Last Visit
- **MTB**: Molecular Tumour Board
- **NGS**: Next Generation Sequencing
- **PROMs**: Patient Reported Outcome Measures
- **REDCap**: Research Electronic Data Capture
- **SPHN**: Swiss Personalized Health Network

## Declarations

## Data Availability

All data produced in the present study will be available upon reasonable request to the authors.

## Acknowledgements

We would like to thank all study coordinators involved in LIQPLAT. We also extend our gratitude to Prof. Matthias Briel for his valuable input to the trial design and his direct supervision of JMS throughout the study development process.

## Authors’ contributions {31b}

1. Conceptualization: AMS, BK, JMS, LGH, PJ
2. Data Curation: ./.
3. Formal Analysis: ./.
4. Funding Acquisition: AMS, BK, JMS
5. Investigation (trial-ongoing): AI, AMS, BK, CCW, JMS, LH, MB, MSM
6. Methodology: AMS, BK, JMS, LGH, PJ
7. Project Administration: AMS, BK, JH, JMS
8. Resources: BM
9. Software: BK, JMS
10. Supervision: BK
11. Validation: ./.
12. Visualization: JMS
13. Writing – Original Draft: AMS, JMS, JP
14. Writing – Review & Editing: all authors

## Availability of data and materials {29}

All data produced in the present study will be available upon reasonable request to the authors.

## Ethics approval and consent to participate {24}

The Ethikkommission Nordwest-und Zentralschweiz (EKNZ) reviewed and approved the protocol (BASEC 2024-00358). Written, informed consent to participate will be obtained from all participants.

## Consent for publication {32}

n/a. The manuscript does not contain any individual person’s data in any form.

